# Signature of the State measures on the COVID-19 Pandemic in China, Italy, and USA

**DOI:** 10.1101/2020.04.08.20057489

**Authors:** Farhan Saif

## Abstract

We show the dynamics of COVID-19 outbreak in Italy and USA, in comparison with China, and the early response of the countries. Our mathematical techniques makes it possible to calculate the rate of growth of the cases efficiently, and provides a good understanding of future trends in Italy and USA. The evolution of the real time data makes it possible to analyse the suitability of steps taken to eradicate the pandemic by the countries. We compare the day to day development of the coronavirus cases in Italy and USA, that keeping in view the population pyramid and the population density, leads us to understand possible difference in the number of effected population.

## a. Introduction

On December 27, 2019, a hospital in Wuhan, capital of Hubei province, reported the first mysterious, suspected pneumonia cases to the center of disease control (CDC) in the capital. On January 8, 2020 a new coronavirus was identified as the cause of pneumonia that spread all over China in next three weeks. Since then, the novel coronavirus, COVID-19, spread across the globe in the next month. In March the World Health Organization declared it to be a pandemic, that in the absence of a vaccine became a challenge for mankind with a tendency to grow exponentially if no measure is taken to prevent it [**?**]. The incubation period of COVID-19 can last for two weeks or longer [2]. During the period of latent infection, the disease may still be infectious [3]. The virus can spread from person to person through respiratory droplets and close contact [4].

Employing social distancing, following patient history case by case, and locking down the province of Hubei, and de-facto quarantining the other provinces by restricting the movement of people China controlled the COVID-19 spread in thirty five days successfully with minimum number of deaths. On the other hand similar out break in Iran and Italy remained uncontrolled after forty five and sixty days, respectively, with forty five thousands in Iran and one hundred and five thousands people in Italy tested positive as on March 31. In this paper based on mathematical modelling and reported previous evolution in China, we predict the future dynamics of the COVID-19 outbreak in the second quarter of 2020 in Italy and USA. The lack of testing facilities and sanctions are restricting the authenticity of data from Iran, for the reason we are not including it in the present publication. For our study related to China we have considered the first twenty one days data that is from January 16, till February 05, whereas we have excluded the epicentre province of Hubei, which had a significant issue of under reporting at the early stage and also data inconsistency due to a change of classification guidelines. For Italy and USA we have considered the data from February 01, till April 06.

Various mathematical model to analyse the growth of an epidemic are employed, that include SEIR model [5], generalized SEIR model [6] and neural network based quarantine control model [7]. Moreover mathematical analysis of pandemics based on exponential growth model [8–11] and generalized growth model [12–22] have been reported. In this contribution, we have developed discrete generalized growth model (DG^2^M), the discrete generalized logistic model (DGLM), and discrete generalized Richard model (DGRM). These models have their limitations and are only applicable to some stages, however they provide more accurate measure of the growth in the outbreak. Based on our numerical results we find excellent agreement between the analytical results and the real-time data of coronavirus positively tested cases. The top-down methods have the advantage to study future scenario of the outbreak, provide lower and upper bounds of our predictions, the rate of growth in stages and provide informative implications of the measures taken to control and eradicate the pandemic.

The manuscript has the following layout: in section II we present the mathematical model for our study, in section III we explain the real-time data of positively tested COVID-19 patients with the numerical results based on our models. In section IV we present our estimates and recommendations based on the work.

## b. Mathematical Model

The generalized Richard Model [23], that is

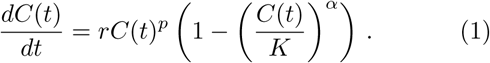

is used in explaining previous epidemics. It is the extension of the original Richards growth model [24] which explain the pandemics [25–29]. Here, *r* is the growth rate at the early stage, and *K* is the final epidemic size. Moreover *p* is a parameter that allows the model to capture different growth profiles. The exponent *α* measures the deviation from the dynamics of the simple logistic curve. The equation (1) corresponds to the original Richards model for *p* = 1, that is

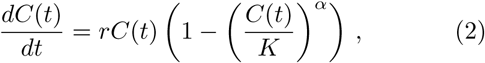

and reduces to the generalized logistic model for *α* = 1 and *p* = 1.

Keeping in view the development of data in terms of the positive tests, variations, and deaths on every next day we may consider the change in *C* over a unit interval of time. We express the rate of change of *C*, that is *Ċ* (*t*), in difference form as 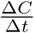. Here Δ*C* = *C*_*n*+1_ − *C*_*n*_ is the change in positively tested patients. Thus we reshape the equation (1) in the discrete form as

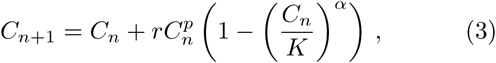

where *C*_*n*_ is the number of positively tested patients on *n*th day. The discrete generalized Richard model (DGRM) provides an effective way to monitor the pandemic in the form of a mapping, thus connecting the data on *n*th day with (*n* + 1)th day. The stable points of the discrete generalized Richards model given in equation (3) are *C*_*n*_ = 0, that implies no positively tested patients, and *C*_*n*_ = *K*. The stable points are the same as obtained by the generalized Richards model.

## c. Numerical results and real-time data

We write the discrete generalized growth model (DG^2^M), as

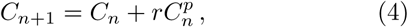

which provides exponential growth for *p* = 1, that is 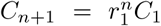, that relates the number of positively tested patients on the first day with the number of postively tested patients after a number of *n* days has passed, here *r*_1_ = 1 + *r*. Hence for the parameter *p* = 0 we find linear increase with respect to *n* as *C*_*n*+1_ = *C*_1_ + *nr*, and sub-exponential growth for 0 < *p* < 1.

The outbreak is discussed in the initial phase in the three countries, that is China, Italy and USA. In the presence of an almost consistent growth of young population and large old population, the population pyramid in the three countries has close resemblance, as shown in figure **??**. The population between 15 and 65 years is 70%, 64% and 63%, whereas above 65 years of age in China, Italy and USA is, respectively, 9.55%, 22.41% and 14.79%. However the pandemic has effected the three countries in diversely different pattern hinting at the preparedness of the countries to tackle such natural disasters as we show in the following discussion.

In order to make a relation between the number of positively tested cases reported every day with the increase in the number of these cases per day, we develop double log plot, shown in figure 1. We compare the data of the patients with the discrete generalized growth model given in equation (4). Taking the logrythm of the equation (4), we write the corresponding relation for the logrythm of the increase in the number of patients log Δ*C*_*n*_ as

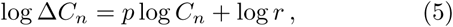

which is the equation of a straight line, in (log Δ*C*_*n*_, log *C*_*n*_) plane. Here *p* expresses the slope of the straight line.

**FIG. 1.**
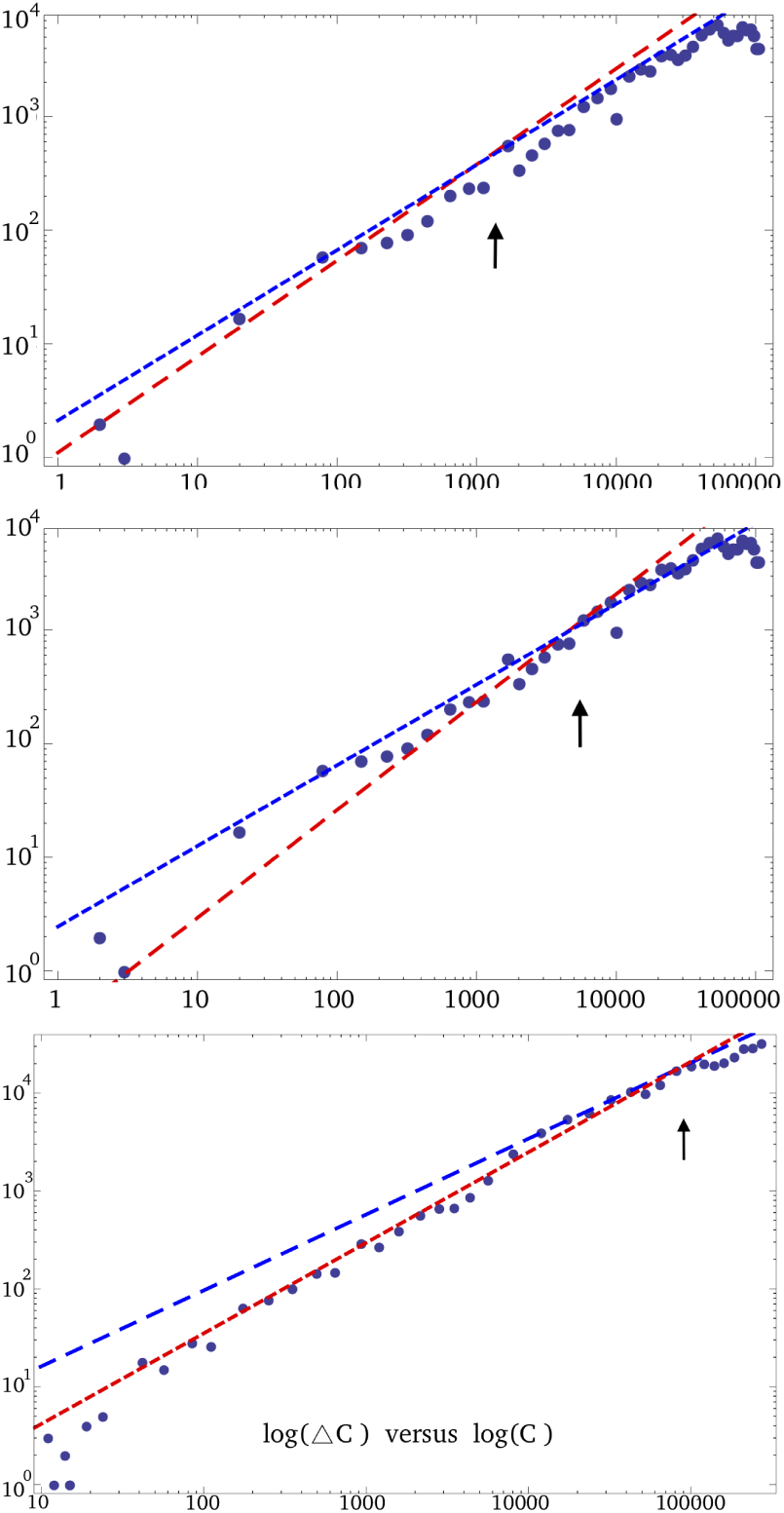
We show log-log plot between the number of positive cases and the day-to-day increase in number in the alphabetical order: In the upper panel we show the real-time data of patients in China showing logrythm of the number of increase in the positively tested COVID-19 patients as a function of logrythm of positively tested patients. We compare mathematical plots obtained by DG^2^M shown in red dashed-line for *r* = 1.105 and *p* = 0.845, whereas the blue dashed-line is plotted by using DGRM for *r* = 2.117, *p* = 0.75, *α* = 1.3, and *K* = 60, 000. In the middle panel we show real-time data for Italy. We compare the mathematical plots obtained by DG^2^M model shown in red dashed-line for *r* = 2.46 and *p* = 0.72, whereas the line plotted by means of DGRM has *r* = 0.33, *p* = 0.95, *α* = 1, and *K* = 155, 000; In the lower panel we show the comparison for USA. Here we have the data obtained by DG^2^M shown in red dashed-line for *r* = 0.202 and *p* = 1.115, whereas the blue dashed-line plotted by employing DGRM for *r* = 2.718, *p* = 0.775, *α* = 1.4, and *K* = 950, 000.

Interestingly a comparison of the real time data of the positively tested patients with equation (5) makes it appear in two sub-groups: First sub-group of the data makes a straight line that describes the evolution of COVID-19 pandemic in its early days. The outbreak follows a certain rate *r* of its spread that define the corresponding straight line as expressed in equation (5). The parameter *p* describes the spread of the outbreak, it is sub-exponential for 0 < *p* < 1, exponential for *p* = 1, and faster than exponential for *p* > 1. It appears in figure 1 as a red line for the three different countries, that is China, Italy and USA respectively; Second, the next stage of the outbreak makes another straight line as shown by blue line in figure 1. The point of intersection of the two lines, indicated by vertical arrows in figure 1, corresponds to the preventive measures adopted by the respective country. For example, the early intersection of the two lines in case of China indicates a general lock down in most of the provinces of China on January 26,2020, as soon as the reported cases were around one thousands. Whereas in the case of Italy it happened after an order of magnitude increase in number that is around 10000, as appears in figure 1. In case of USA we find the intersection point appears even farther.

We obtain the mathematical results by the discrete generalized growth model, discrete generalized logistic growth model and the discrete generalized Richard model by using the *r* and *p* values obtained from above discussion for China, Italy and USA. The obtained results are compared with the real time data of COVID-19 patients obtained from these countries, as shown in figure 2.

**FIG. 2.**
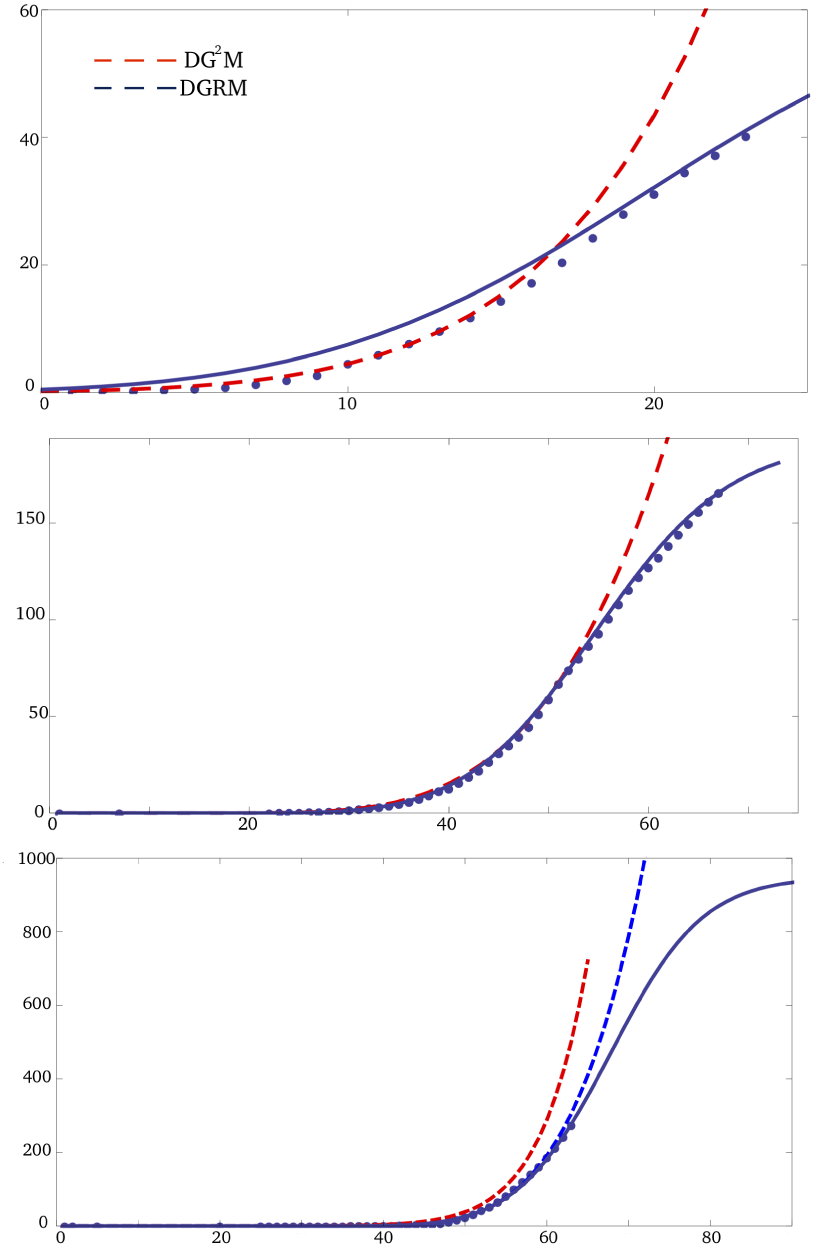
We plot the number (in thousands) of positively tested cases versus the number of days, in the alphabetical order for China, Italy and USA. The description of the red dashed-line and blue dashed-line, and the values of the parameters is the same as given in figure 1. The blue dashed line in lower panel dedicated to USA data shows the pandemic dynamics following DG^2^M, whereas the blue line expresses the COVID-19 pandemic dynamics following DGRM.

The plots show that the discrete generalized growth model (DG^2^M) show the gradual increase that matches very good with the real time data of positively tested patients in the three countries. The preventive measures taken in China show a gradual deviation from the DG^2^M model, whereas it is taken over by the DGRM. In case of Italy the gradual deviation of real-time data is also seen, reflecting a slow down of the outbreak, that indicates a positive effect of the steps taken by the government of Italy, such as strict lock down, intercity travel and closure of education sector, and unnecessary travels. In case of USA, the data is showing a deviation from DG^2^M, however it is not significantly close of DGRM. The blue dashed-line indicates the future growth of the coronavirus pandemic following DG^2^M model showing an uncontrolled increase. Whereas the blue line shows the future trend of the pandemic following DGRM model. This implies that to eradicate the COVID-19 successfully it is required to take further strict measures.

The logrhythmic evolution, in figure 3, expresses the size of the outbreak in China, Italy and USA. As the estimated size of outbreak agrees well with the reported data of China and Italy, it is expected that in USA in the absence of further strict measures the coronavirus pandemic may effect around a million people in next fifteen days, almost 0.3% of USA population in another five days and before we see a slowing down effect.

**FIG. 3.**
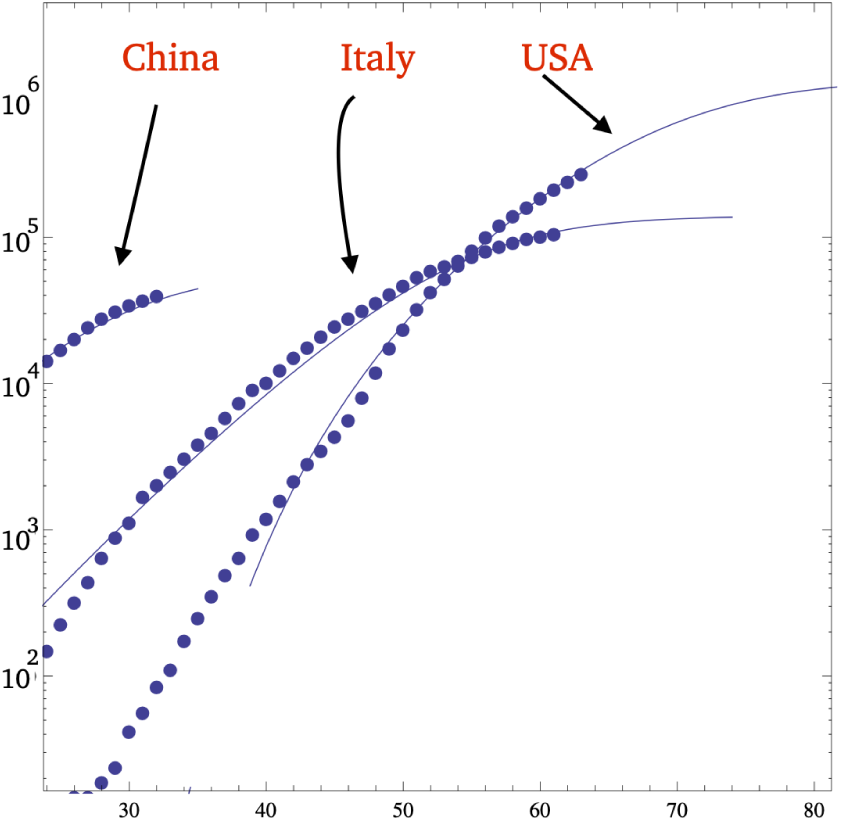
Along the vertical axis we show the logrythm of number of positively tested cases of COVID-19, and the number of days are along the horizontal axis. The blue dashed-lines are obtained by DGRM whereas the blue dots represent the real-time data. The values of the parameters are the same as given in figure 1.

## d. Conclusions

We show that an early response from Italy and USA was necessary to restrict the wide spread of the pandemic. In comparison with the COVID-19 early evolution in China we provide quantitative analysis of the outbreak in Italy and USA. We mathematically identify the transition in the evolution of the pandemic as a function of measure taken by respective governments by classification of corresponding data in stages which are defined by mathematical parameters. We provide rate of growth at every stage, and measure of the nature of expansion of pandemic. We predict a high risk in USA, where the number of positively tested coronavirus cases may reach one million in the absence of more strict measures in next fifteen days, and in next twenty days 0.3% USA population may get infected in a positive

## Data Availability

The data of the number of patients in every country is taken from wikipedia
https://en.wikipedia.org/wiki/2020_coronavirus_pandemic_in_the_United_States
https://en.wikipedia.org/wiki/2020_coronavirus_pandemic_in_Italy
https://en.wikipedia.org/wiki/2020_coronavirus_pandemic_in_china

